# A genetic and transcriptomic assessment of the *KTN1* gene in Parkinson’s disease risk

**DOI:** 10.1101/2021.03.08.21252688

**Authors:** Anni Moore, Sara Bandres-Ciga, Cornelis Blauwendraat, Monica Diez-Fairen, on behalf of the International Parkinson’s Disease Genomics Consortium (IPDGC)

**Author notes:** **Corresponding author:** Dr. Monica Diez-Fairen,. University Hospital MútuaTerrassa, Sant Antoni 19, Terrassa 08221, Barcelona, Spain.

## Abstract

Parkinson’s disease (PD) is a progressive neurological disorder caused by both genetic and environmental factors. A recent finding has suggested an association between *KTN1* genetic variants and changes in its expression in the putamen and substantia nigra brain regions and an increased risk for PD. Here, we examine the link between PD susceptibility and *KTN1* using individual-level genotyping data and summary statistics from the most recent genome-wide association studies (GWAS) for PD risk and age at onset from the International Parkinson’s Disease Genomics Consortium (IPDGC), as well as whole-genome sequencing data from the Accelerating Medicines Partnership Parkinson’s disease (AMP-PD) initiative. To investigate the potential effect of changes in *KTN1* expression on PD compared to healthy individuals, we further assess publicly available expression quantitative trait loci (eQTL) results from GTEx v8 and BRAINEAC and transcriptomics data from AMP-PD. Overall, we found no genetic associations between *KTN1* and PD in our cohorts but found potential evidence of differences in mRNA expression, which needs to be further explored.

## Introduction

Parkinson’s disease (PD) is a neurodegenerative disorder caused by both genetic and environmental risk factors. Its neuropathological hallmark is the loss of midbrain dopaminergic neurons in the substantia nigra pars compacta (SNpc) that innervate the basal ganglia, a set of subcortical nuclei that play a central role in motor control, motor learning, executive functions, and emotion regulation ^1^. In PD, the progressive degeneration of these neurons leads to severe dopamine (DA) deficiency in the striatum and the subsequent appearance of the cardinal motor features of the disease, including tremors, rigidity, hypokinesia, and postural imbalance ^2,3^. Several treatments for PD are known to target the SNpc as well as the putamen such as the pharmacological substitution of striatal dopamine using L-DOPA, the immediate biosynthetic precursor of DA ^4^, and surgical interventions like deep brain stimulation for those patients developing intractable L-DOPA-related motor complications ^5^.

Recently, Mao *and colleagues* reported nominal associations between common intergenic variants downstream and upstream of the *KTN1* gene and PD risk, as well as an increased *KTN1* mRNA expression in the putamen and SNpc, which resulted in a compensatory increase in the gray matter volumes (GMV) of these two brain regions in PD patients, suggesting that *KTN1* may play a functional role in the development of PD ^6^. *KTN1* encodes the integral membrane protein kinectin 1, a receptor that is involved in organelle transport within the cell by allowing vesicle binding to kinesin ^7^. Impairment of these molecular motors has been increasingly linked to neurological diseases with a subcortical component, such as PD ^8^. In addition, rs945270, an intergenic locus downstream of *KTN1*, has been associated with larger putamen GMV and an increased *KTN1* expression in the frontal cortex and putamen brain regions and blood tissue ^9^. More recently, variants in close proximity to *KTN1* were also associated with the volume of the nucleus accumbens, caudate nucleus, and globus pallidus, suggesting that this genomic region may have an important role in determining multiple subcortical brain volumes during development ^10^.

In the study by Mao *et al*, a total of eight intergenic variants near *KTN1* were nominally associated with PD. This included six variants associated with PD across at least two independent cohorts and two other GMV-related variants that were not found in the discovery dataset but nominally associated with PD in one of the two replication cohorts. Of note, none of the reported *KTN1* associated variants were consistently replicated across all cohorts. In addition, six of these risk variants were found to regulate either the mRNA expression in putamen, SNpc or putamen GMV, and *KTN1* was differentially expressed in the SNpc between PD and controls in one cohort. As part of the International Parkinson’s Disease Genomics Consortium (IPDGC) efforts to investigate and replicate reported risk factors for PD, we sought to assess the role of *KTN1* as a genetic risk factor for PD within large European cohorts. Here, we considered all potential influencers of *KTN1* by including variants in the entire cis-*KTN1* region to account for their potential role in regulating *KTN1* mRNA expression in the putamen and SNpc.

## Methods

### Genotype data

Imputed individual-level genotyping data from genome-wide association studies (GWAS) were obtained from the IPDGC and contained 14,671 PD samples and 17,667 healthy controls, all of European ancestry. All datasets underwent quality control procedures as previously reported ^11,12^. Of note, one of Mao *et al* replication datasets, “*lng_coriell_pd*” (phs001172.v1.p2), is partially included in IPDGC genotype data we used. Data were based on the hg19 genome assembly, expanding the *KTN1* region (chr14: 56,025,790-56,168,244) by 1MB on each end to include the entire cis-*KTN1* region (chr14: 55,025,790-57,168,244), and were extracted with PLINK1.9 ^13^. Then, PLINK1.9 was used to perform a logistic regression adjusted by sex, age, 10 principal components (PCs) to account for population stratification, and cohort to control for chip bias. Variants were annotated using ANNOVAR ^14^.

We further assessed data from the most recent GWAS meta-analyses to date (excluding 23andMe data) for PD risk consisting of 15,056 PD cases, 18,618 UK Biobank proxy-cases (i.e., subjects with a first degree relative with PD) and 449,056 controls ^11^, as well as for age at onset (AAO) comprising 17,996 PD cases^12^. All participants were of European ancestry and details on genotyping quality and imputation are described in the original sources. Summary statistics used in this study are public and available via https://pdgenetics.org.

### Whole-genome sequencing data

Whole-genome sequencing (WGS) data were retrieved from the Accelerating Medicines Partnership - Parkinson’s Disease Initiative (AMP-PD; www.amp-pd.org) which contains three cohorts including Parkinson’s Progression Markers Initiative (PPMI; https://www.ppmi-info.org/), Parkinson’s Disease Biomarkers Program (PDBP; https://pdbp.ninds.nih.gov/), and BioFIND (https://biofind.loni.usc.edu/). In total, the set contained 1,647 PD patients and 1,050 healthy controls of European ancestry, with an average AAO of 59.7 years in cases and an average age of 60.1 years in controls. More detailed cohort characteristics, as well as quality control procedures, are further described in https://amp-pd.org/whole-genome-data. Again we used the 1MB expanded cis-*KTN1* region, (chr14: 54,559,072-56,701,526) based on the hg38 genome assembly, and extracted it with PLINK1.9. Results were annotated with ANNOVAR. Association analyses of *KTN1* variants and risk for PD were done using single-variant score tests in RVTESTS package v2.1.0 ^15^, with a minimum allele count (MAC) threshold of 3 applied, and adjusted by sex, age, 10 PCs, and origin cohort to account for underlying variables.

To assess the cumulative effect of multiple rare variants on the risk for PD, we performed gene-based burden analyses on WGS data. Including all variants within the region, a minimum allele count (MAC) threshold of 1 was applied, and variants with a minor allele frequency (MAF) < 1% were filtered out before running burden tests (SKAT, sequence Kernel association test, and SKAT-O, optimized SKAT) with RVTESTS package v2.1.0 following default parameters ^15^. Age, gender, 10 PCs, and the dataset were again accounted for covariates. All code used in this study can be found at https://github.com/ipdgc/IPDGC-Trainees/blob/master/KTN1.md.

### Expression Quantitative Trait Loci (eQTL)

We chose to investigate eQTL values of all genes found within the cis-*KTN1* region to account for potential effects on expression by any variants within the region. Variant eQTL results of all genes were obtained from the GTEx Analysis Release V8 (dbGaP Accession phs000424.v8.p2) based on analyses from WGS genotypes and RNA-seq expression data from putamen and SNpc tissue samples. GTEx subjects are predominantly of European ancestry (84.6%) but do contain other ancestries as well. Putamen eQTL results were calculated from 170 total samples with genotype and expression data. SNpc eQTL values were calculated from 114 patient samples ^16^. We also obtained values from BRAINEAC (UKBEC) ^17^, which used genotype data and reads per kilobase per million (RPKM) normalized and covariate-adjusted expression data from 117 post-mortem putamen samples of European ancestry to calculate eQTL values ^18^.

### Transcriptomics data

Whole blood time progression gene expression data (gencode v29) from 0-24 months was accessed from AMP-PD. We used the PPMI, PDBP, and BioFIND cohorts at the baseline, 0 month time point (note BioFIND ‘Baseline’ is at ‘0.5’ month), including a total of 1,886 cases and 1,285 control samples. Samples with sex mismatch were excluded. Raw mRNA expression of *KTN1* in all datasets can be seen in **Figure 1**. Expression data were quantified as transcripts per kilobase million (TPM) and quantile normalized before *KTN1* values were extracted using Ensembl gene id ENSG00000126777.17 (**Figure 2**). Using the scikit-learn Python packages ^19^, residuals were calculated using linear regression, and the data were adjusted using age, sex, race, 20 genotype PCs, cohort, and missingness rate as covariates (**Figure 2**). To test for significant differences in expression between the cases and controls, we performed a t-test with the residuals. We also used an additional dataset taken from the 24-month time point in the PDBP and PPMI cohorts of 841 cases and 386 controls to confirm our findings.

**Figure 1.**
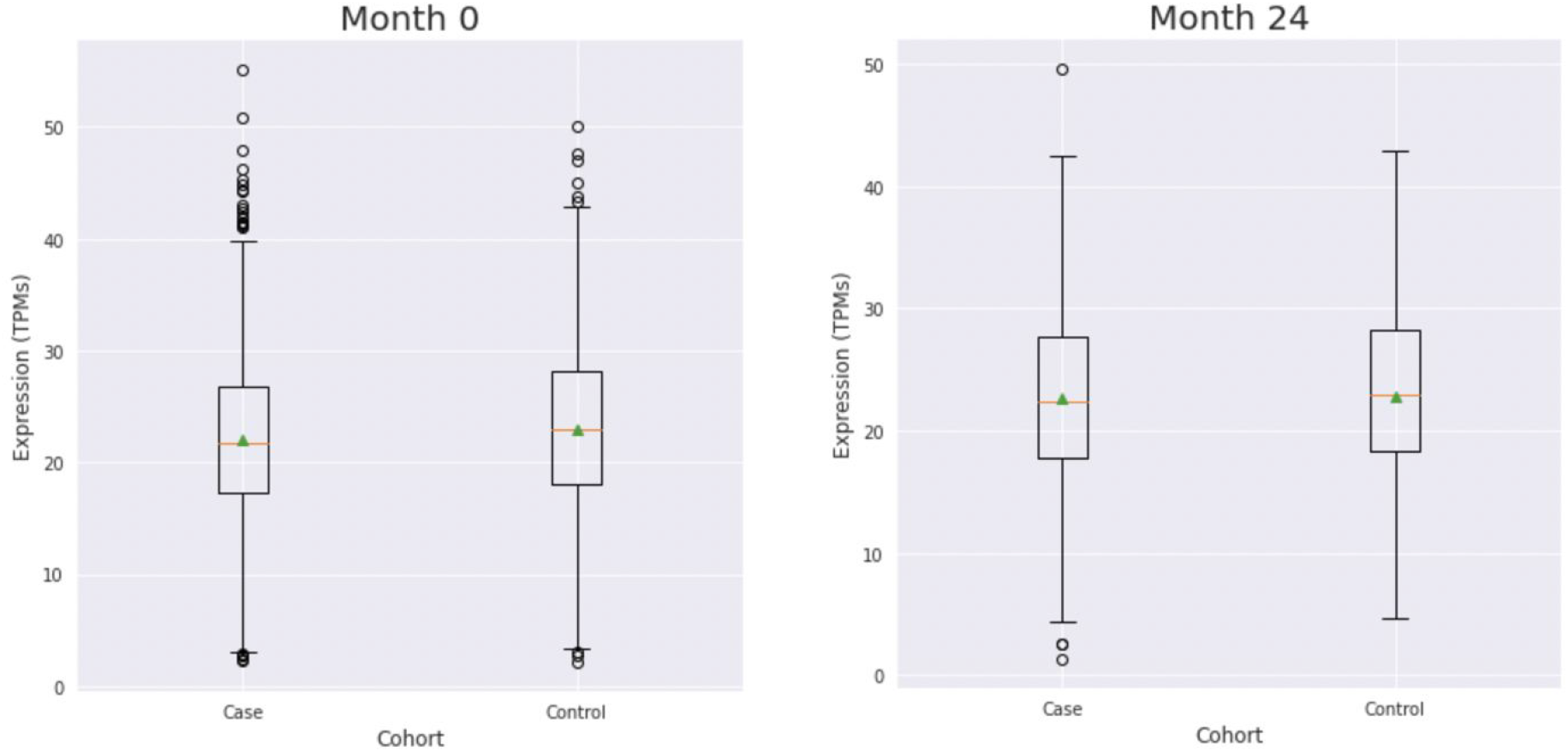
Raw *KTN1* mRNA expression - expressed in transcripts per kilobase million (TPMs) - by cohort in both tested AMP-PD expression datasets.

**Figure 2.**
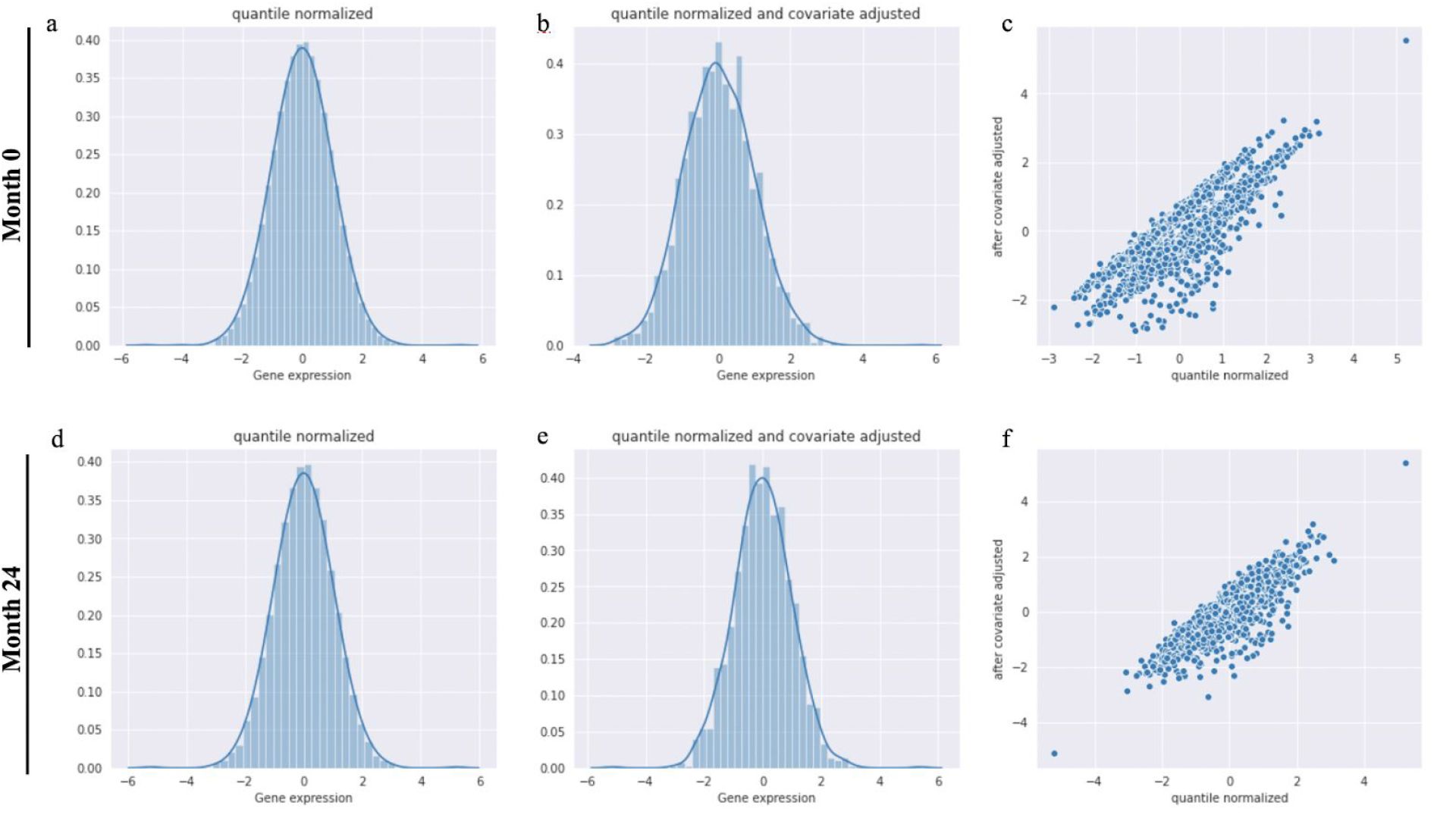
KTN1 mRNA expression processing. **a**. Distribution of all gene expression after quantile normalization at Month 0. **b**. Distribution of all gene expression after quantile normalization and covariate adjustment at Month 0. **c**. Linear regression of quantile normalized values versus quantile normalized and covariate adjusted at Month 0. **d**. Distribution of all gene expression after quantile normalization at Month 24. **e**. Distribution of all gene expression after quantile normalization and covariate adjustment at Month 24. **f**. Linear regression of quantile normalized values versus quantile normalized and covariate adjusted at Month 24.

### Ethics Statement

We confirm that our study does not need ethical approval from the institutions involved since it involves information freely available in the public domain, such as the analysis of open-source datasets, where the data was properly anonymized and informed consent was obtained at the time of original data collection.

## Results

### Association between KTN1 variants and PD risk

Using imputed genotype data from GWAS, we identified a total of 6,188 variants present in the cis-*KTN1* region (Supplementary Table 1). We performed a case-control association analysis using logistic regression which showed no variants significantly associated with PD (p<5·10^−8^). Among the 6,188 variants found, 35 were exonic, with 19 of them being synonymous and 16 nonsynonymous variants. The remaining 6,153 non-coding variants included 2,331 intronic, 54 in the 3’-untranslated region (3’-UTR) and 8 in the 5’-untranslated region (5’-UTR) (Supplementary Table 1). All variants found by Mao *and colleagues* were noncoding, including five variants (rs12880292, rs8017172, rs17253792, rs945270, rs1188184) that were intergenic, two in non-coding exonic regions (rs7142488, rs4144657), and one that was downstream (rs7157819). We further analyzed summary statistics from the above-mentioned risk and AAO PD GWAS meta-analyses. No evidence for an association between *KTN1* common genetic variation and PD was detected (p<5·10^−8^) for either PD risk or AAO (**Figure 3**).

**Figure 3.**
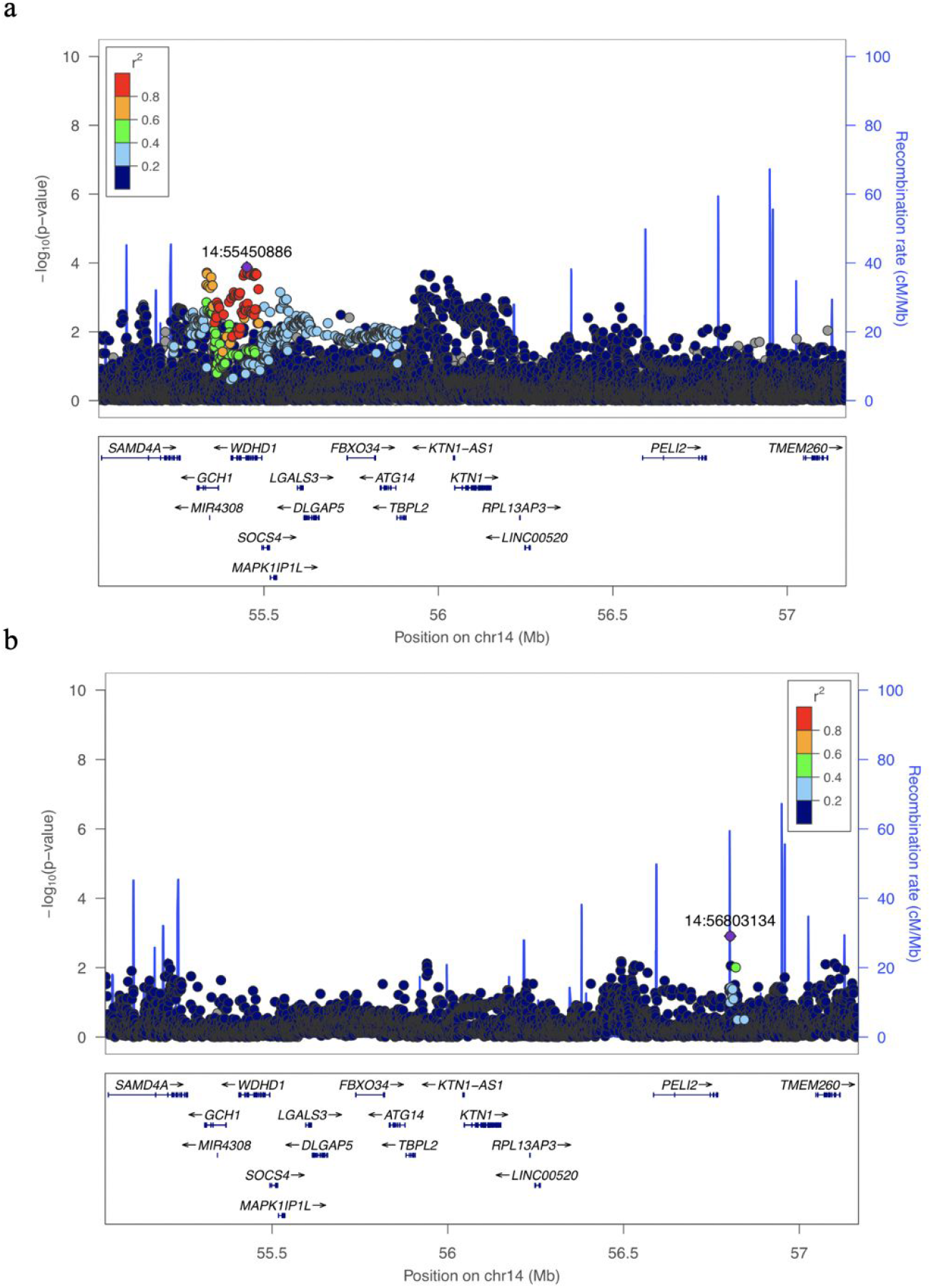
*KTN1* locus zoom plots for PD risk and age at onset (AAO) plotted as the base-pair position of each variant (x-axis) versus the -log10(p-value) of variants on or near *KTN1* (y-axis). Variants are colored by their R2 linkage disequilibrium value, which is relative to the variant with the lowest p-value, shown in purple. **a**. Plotting of p-values from PD risk GWAS meta-analysis of cis-*KTN1* region, excluding 23andMe data ^11^. **b**. Plotting of p-values from PD AAO GWAS meta-analysis of cis-*KTN1* region, excluding 23andMe data ^12^.

Similarly, using WGS data, we identified 17,382 variants in the cis-*KTN1* region (Supplementary Table 2), 337 of which were coding including 137 synonymous and 193 non-synonymous variants. Upon performing a case-control analysis using a single score variant test, no significant difference was determined (p<5·10^−8^) in the frequency of *KTN1* variants between PD and healthy individuals. Gene-based burden analyses showed no enrichment of rare variants (p<5·10^−8^) in cases compared to controls (Supplementary Table 3).

Of note, the original eight variants reported by Mao *and colleagues* were not significantly associated with PD (p<5·10^−8^) in our genotyping nor sequencing datasets (**Table 1**).

**Table 1.** Variants from Mao *and colleagues* in all datasets.

| Position<br>(hg19) | Position<br>(hg38) | rsID | Ref | Alt | Frequency |  |  |  |  | P-values |  |  |
| --- | --- | --- | --- | --- | --- | --- | --- | --- | --- | --- | --- | --- |
|  |  |  |  |  | IPDGC<br>Case | IPDGC<br>Control | GWAS<br>Summary<br>Statistics* | AMP-PD | gnomAD<br>NFE | IPDGC | GWAS<br>Summary<br>Statistics* | AMP-PD |
| 14:55952864 | 14:55486146 | rs12880292 | G | A | 0.3059 | 0.3250 | 0.321 | 0.310 | 0.338 | $1.90 \cdot 10^{-7}$ | 0.001 | 0.649 |
| 14:56199048 | 14:55732330 | rs8017172 | G | A | 0.4033 | 0.4182 | 0.414 | 0.422 | 0.418 | $1.39 \cdot 10^{-4}$ | 0.011 | 0.045 |
| 14:56205030 | 14:55738312 | rs17253792 | T | C | 0.0873 | 0.0892 | 0.905 | 0.085 | 0.079 | 0.384 | 0.343 | 0.108 |
| 14:56200473 | 14:55733755 | rs945270 | C | G | 0.4041 | 0.4185 | 0.585 | 0.423 | 0.418 | $2.06 \cdot 10^{-4}$ | 0.015 | 0.050 |
| 14:56247593 | 14:55780875 | rs7157819 | C | T | 0.2989 | 0.2974 | 0.303 | 0.295 | 0.290 | 0.691 | 0.524 | 0.859 |
| 14:56248010 | 14:55781292 | rs7142488 | T | C | 0.2997 | 0.2987 | 0.697 | 0.296 | 0.291 | 0.784 | 0.452 | 0.852 |
| 14:56248455 | 14:55781737 | rs4144657 | G | A | 0.0657 | 0.0688 | 0.073 | 0.069 | 0.075 | 0.115 | 0.055 | 0.780 |
| 14:56442412 | 14:55975694 | rs1188184 | G | A | 0.3649 | 0.3655 | 0.371 | 0.380 | 0.608 | 0.872 | 0.837 | 0.469 |

Key: AMP-PD, Accelerating Medicines Partnership Parkinson's disease; IPDGC, International Parkinson's Disease Genomics Consortium; GWAS, Genome Wide Association Study; NFE, Non-Finnish Europeans.
\*Summary statistics frequency from Nalls et al. 2019 <sup>11</sup>

### KTN1 and mRNA expression

To test the potential regulatory effect of cis 1MB region for *KTN1* risk variants affecting the level of expression of *KTN1* mRNA in the putamen and substantia nigra brain regions, we first considered cis-eQTL results from the GTExv8 cohort. 18 genes were found within the cis-*KTN1* region including *DLGAP5, KTN1-AS1, LGALS3, SOCS4, SAMD4A, MIR4308, TBPL2, KTN1, PRL13AP3, LINC00520, PELI2, LOC101927690, FBXO34, MAPK1IP1L, TMEM260, GCH1, WDHD1*, and *ATG14*. After filtering results from all genes by p-value for both brain tissue types, we only found two significant values for the gene *ATG14* in the putamen. Both variants, rs112424815 (p=6.10e^-8^) and rs79509284 (p=6.90e^-8^) fall within our cis-*KTN1* region. No significant variants were found in the substantia nigra. We also consulted eQTL results from BRAINEAC derived from UKBEC, which showed no significance from any gene variants for putamen or substantia nigra tissue.

Using mRNA gene expression data from whole blood, we assessed the difference in *KTN1* expression between 1,886 cases and 1,285 control samples from AMP-PD. We performed a t-test between normalized, covariate-adjusted *KTN1* expression in cases *vs* controls, and found a significant difference in *KTN1* mRNA expression in whole blood (p=0.0012) at the 0 month time point (**Table 2**). However, when sub-cohorts PPMI and PDBP were separated within the dataset and run individually to test if any single sub-cohort was driving overall significance, neither cohort indicated a significant difference between *KTN1* expression in cases vs controls (Supplementary Table 4). The initial test was also not consistent in additional whole blood expression data from a follow-up visit at 24 months from AMP-PD for 841 cases and 386 controls. We calculated a p-value of 0.8112 for the t-test, indicating a non-significant difference (**Table 2**). Therefore we cannot conclude that a significant difference exists in *KTN1* mRNA expression between PD patients and controls in whole blood, and further analyses should be done.

**Table 2.**
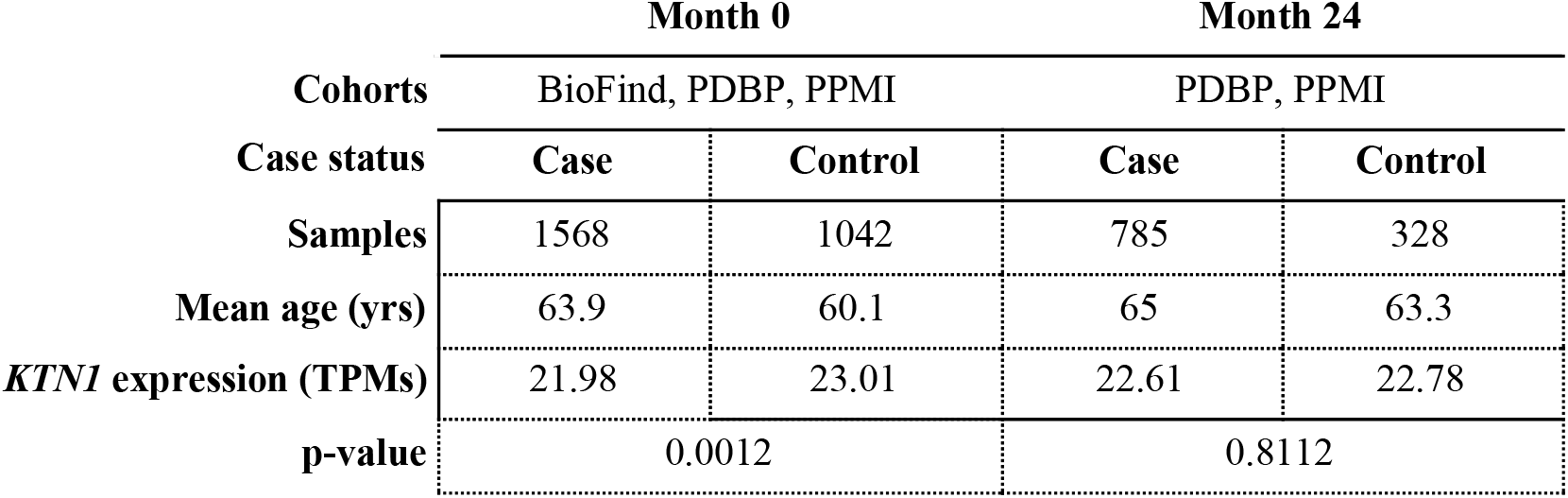
T-test in AMP-PD whole blood mRNA expression.

## Discussion

As a substantial proportion of PD risk is driven by genetic factors, efforts are continually being made to determine new genes that may be linked to the disease. While genes like *LRRK2, SNCA*, and *GBA* are well-known to have a substantial impact on one’s likelihood of developing PD, they only explain a small fraction of PD heritability, suggesting that additional genes may harbor rare and common variation associated with the disease ^11,20^. In addition, in most cases, it is still unknown how the identified genetic risk variants disrupt molecular and cellular processes underlying the pathobiology of the disease. With many of the known PD-related mutations affecting genes involved in intracellular transport systems ^21^, *KTN1* may be a relevant gene to investigate given its role in organelle transport and vesicle binding ^7^.

In our study, we were not able to replicate the findings by Mao *and colleagues* and cannot conclude that the *KTN1* gene is significantly implicated in PD etiology. Using large European cohorts, we were not able to find any variants in the cis-*KTN1* region significantly associated with PD when compared to healthy individuals. Analysis of summary statistics from the most recent and largest GWAS meta-analyses to date for PD risk and AAO also showed no significant variants (p<5·10^−8^). We were not able to find a higher frequency of *KTN1* rare variants among PD patients. All eight *KTN1* variants found by Mao *et al*. were found outside the *KTN1* gene, one upstream and seven downstream, which we covered by considering the entire gene cis-region. None of the variants were significantly associated with PD in our analyses. Therefore we could not find genetic evidence of any association between *KTN1*-related variants and PD. We recognize that these findings only represent patients of European ancestry and we cannot exclude association between *KTN1* and PD in other populations.

Despite not finding genetic evidence, we continued to consider any potential effects of *KTN1* expression on PD risk. We consulted public databases, GTEx and BRAINEAC, consisting of hundreds of genotype and expression samples from the SNpc and putamen brain regions. Of note, one of the 18 genes within the region, *GCH1*, has been previously found to be associated with PD ^11^. *GCH1* encodes the enzyme GTP cyclohydrolase 1, which is necessary for dopamine synthesis in nigrostriatal cells ^22,23^ and dopamine deficiency in the nigrostriatal region is a core biological feature of PD ^24^. However, we found no significant eQTL values for *GCH1*. We were only able to locate two eQTL values of significance within the cis*-KTN1* gene region for the putamen - both variants affecting the expression of the gene *ATG14*. No eQTL values were found to be significant in SNpc tissue for any gene within the region. *ATG14* function is required for both basal and inducible autophagy but also determines the localization of the autophagy-specific PI3-kinase complex and contributes to autophagosome formation ^25^. Interestingly, there is genetic, pathological, and experimental evidence suggesting an important role for autophagy impairment in PD ^26,27^.

To test for differences in *KTN1* expression we used transcriptomic data from large European cohorts to perform a test on overall expression in whole blood. We did initially note a significant difference (p=0.0012) in *KTN1* mRNA expression in PD patients versus control samples, however upon replication in an additional dataset, we could not reproduce significant results, warranting further investigation in this area. Unfortunately, we only had access to expression data from whole blood and we acknowledge that expression may be different in brain tissues. However, it has been shown that blood samples act as reliable alternatives for expression when brain tissues are not available ^28^. Using blood also allowed us to increase the overall number of samples considerably, contrary to brain samples which are often harder to acquire.

*KTN1* has been previously associated with other neurological disorders, especially impacting putamen volume and structure, providing further evidence that it potentially plays a significant role in the brain. A previously mentioned study by Hilbar and colleagues found that a specific variant of *KTN1*, rs945270, was significantly linked to *KTN1* expression in blood, frontal cortex, and putamen tissues, ultimately resulting in a larger putamen volume ^9^, though we were not able to conclude a similar association in our blood datasets. *KTN1* has also been implicated in alcohol and drug dependency, where the putamen GMV has been shown to be decreased in alcohol-dependent patients ^29^ but increased in stimulant drug-dependent patients ^30,31^. Luo *et al* found significant *KTN1* variants that increased risk for substance abuse disorders in Europeans, as well as increased *KTN1* mRNA expression levels in the putamen and putamen GMVs that led them to conclude a positive relationship existed among variants of *KTN1* and substance use disorders risk ^32^. Thus, despite our inability to conclude a casual increased risk for PD, *KTN1* remains of interest in brain disorders, especially those affecting the putamen area.

In conclusion, based on the current data presented, our analyses do not support a role for *KTN1* variants in PD genetic susceptibility. However, differences in *KTN1* mRNA expression between PD patients and healthy individuals should be further studied. The vast majority of our datasets are of European ancestry, therefore we cannot rule out a potential role of *KTN1* rare variants in PD among other populations. Additional large-scale familial and case-control studies in non-European ancestry populations are necessary to further evaluate the role of *KTN1* in PD etiology.

## Data Availability

The summary statistics from PD meta-analyses that support the findings of this study are available from IPDGC consortium website, https://pdgenetics.org/resources. The whole genome sequencing and transcriptomics data that support the findings of this study are available from AMP-PD website, https://amp-pd.org/. Individual-level genotyping data that support the findings of this study are available from the corresponding author upon reasonable request. All code used in this study is publicly available at https://github.com/ipdgc/IPDGC-Trainees/blob/master/KTN1.md.

https://amp-pd.org/

https://github.com/ipdgc/IPDGC-Trainees/blob/master/KTN1.md

## Acknowledgments

We would like to thank all of the subjects who donated their time and biological samples to be a part of this study. We also would like to thank all members of the International Parkinson’s Disease Genomics Consortium (IPDGC). For a complete overview of members, acknowledgments and funding, please see http://pdgenetics.org/partners. This work was supported in part by the Intramural Research Programs of the National Institute of Neurological Disorders and Stroke (NINDS), the National Institute on Aging (NIA), and the National Institute of Environmental Health Sciences both part of the National Institutes of Health, Department of Health and Human Services; project numbers 1ZIA-NS003154, Z01-AG000949-02 and Z01-ES101986. In addition, this work was supported by the Department of Defense (award W81XWH-09-2-0128), and The Michael J Fox Foundation for Parkinson’s Research. Data used in the preparation of this article were obtained from the AMP PD Knowledge Platform. For up-to-date information on the study, visit https://www.amp-pd.org. AMP PD – a public-private partnership – is managed by the FNIH and funded by Celgene, GSK, the Michael J. Fox Foundation for Parkinson’s Research, the National Institute of Neurological Disorders and Stroke, Pfizer, and Verily. We would like to thank AMP-PD for the publicly available whole-genome sequencing data, including cohorts from the Fox Investigation for New Discovery of Biomarkers (BioFIND), the Parkinson’s Progression Markers Initiative (PPMI), and the Parkinson’s Disease Biomarkers Program (PDBP). This work utilized the computational resources of the NIH HPC Biowulf cluster (http://hpc.nih.gov).

## Competing interests

The authors declare that they have no conflict of interest.

## Author contributions

A.M. contributed to conceptualization and design of the work, formal analysis, curation and interpretation of data, and drafting the manuscript. M.D.F. contributed to conceptualization and design of the work, interpretation of the data, supervision and reviewing the manuscript. S.B.C. and C.B. contributed in reviewing and editing the final manuscript.

## Notes

### Competing Interest Statement

The authors have declared no competing interest.

### Author Declarations

Participants clinical information and genetic samples were obtained with appropriate written consent and local institutional and ethical approval. All necessary patient/participant consent has been obtained and the appropriate institutional forms have been archived. Additionally, our project is covered by all relevant, local IRBs from which the data were derived. All data were de-identified prior to sharing with researchers, and therefore, researchers did not have access to any information on the included subjects. As a result, our study design was reviewed by the NIH Office of IRB Operations, and the reviewing body determined that the research activities did not require IRB approval or review. UK Biobank data were obtained under the application number 33601.

